# Perceived Risk and Distress related to COVID-19: Comparing Healthcare versus non-Healthcare Workers of Pakistan

**DOI:** 10.1101/2020.10.23.20218297

**Authors:** Adeel Abid, Hania Shahzad, Hyder Ali Khan, Suneel Piryani, Areeba Raza Khan, Fauziah Rabbani

## Abstract

**Background:** Healthcare workers (HCWs) find themselves susceptible to contracting COVID-19 or being the source of exposure for their family members. This puts them at a high risk of psychological distress which may compromise patient care. In this study we aim to explore the risk perceptions and psychological distress between HCWs and non-healthcare workers (NHCWs) in Pakistan.

**Methods:** A cross-sectional study was conducted in Pakistan using an online self-administered questionnaire. Respondents were categorized into HCWs (completed or aspiring to complete education in Medicine or allied fields) and NHCWs. HCWs were further categorized into front-line (direct patient care) and back-end HCWs.

**Results:** Data from 1406 respondents (507 HCWs and 899 NHCWs) was analyzed. No significant difference was observed between HCWs and NHCWs’ perception of susceptibility and severity towards COVID-19. Healthcare graduates perceived themselves (66% students vs. 80% graduates, p-value 0.011) and their family (67% students vs. 82% graduates, p-value 0.008) to be more susceptible to COVID-19 than the healthcare students. Frontline HCWs perceived themselves (83% frontline vs. 70% back-end, p-value 0.003) and their family (84% frontline vs. 72% back-end, p-value 0.006) as being more susceptible to COVID-19 than back-end healthcare professionals. Over half of the respondents were anxious (54% HCWs and 55% NHCWs). Female gender, younger age and having COVID-19 related symptoms had a significant effect on the anxiety levels of both HCWs and NHCWs.

**Conclusion:** Frontline HCWs, healthcare students, young people, females and individuals with lower income were at a higher risk of psychological distress due to the pandemic. Government policies should thus be directed at ensuring the mental well-being of frontline HCWs, and improving their satisfaction in order to strengthen health care delivery system.

## Introduction

COVID-19 has grappled the world since its first case was diagnosed in Wuhan, China (1). This has resulted in a global socio-economic crisis and challenged healthcare systems throughout the world. According to the World Health Organization (WHO), this is the worst pandemic the organization has seen in its 75-year long history (2). As of 25^th^ July 2020, Pakistan has officially reported 270,113 confirmed cases with near 5,822 confirmed deaths (3).The pandemic has also taken an economic toll on the population due to the government imposing a nation-wide lockdown from March to May, 2020. According to a Gallup Survey reported in April 2020, 6.9 million households claimed to have reduced the number or size of meals for some family members while nearly 1 in 4 Pakistanis said that they were relying on less preferred and less expensive food items to cover their basic household needs. Furthermore, almost 1 in 5 people in the country say that they had to lean on their savings to cover basic household needs (4). A major concern in the country is the impact of COVID-19 on healthcare workers (HCWs), who are at a high risk during novel disease outbreaks. HCWs around the world are at the forefront in screening, quarantining, and managing actual and suspected COVID-19 patients, creating awareness about risks, and advocating for preventive measures (5). Consequently, fear of contracting the disease is likely to put them under greater stress when compared to the general population.

Experience from severe acute respiratory syndrome (SARS) and H1N1 epidemics underlines that the psychological strain on healthcare professionals, especially those working on the frontlines is significant (6,7). In these stressful times, HCWs are risking high morbidity and mortality due to the nature of their job. Demands of the job may negatively impact the emotional and psychological well-being of those working on the front line. Moreover, as compared to the general public, HCWs face more personal worries such as comparatively greater infection risk to self and others and concerns regarding the well-being of family members. Increased anxiety and depression among frontline healthcare professionals is a common feature in epidemics (8,9). Moreover, there are disparities in terms of psychological impact due to COVID-19 within HCWs depending on their level of patient care. In a study conducted in Singapore the psychological distress, depression, anxiety, and stress experienced by health care workers during the COVID-19 outbreak, frontline nurses had significantly lower vicarious traumatization scores than non–frontline nurses and the general public (10). Another recent study among healthcare professionals in a tertiary infectious disease hospital for COVID-19 in China also revealed a high incidence of anxiety and stress disorders among frontline medical staff, with nurses having a higher incidence of anxiety than doctors (11).

The scale of the current health crisis is a bigger concern in a resource-limited country like Pakistan, where it may significantly compromise the quality of care and health care services. These unprecedented times call for an increased need to gauge the knowledge and protective behaviors of people at different levels of health care services. Unduly high anxiety levels and poor-risk perceptions often result in barriers to healthcare providers’ willingness and ability to work and constitute an important component in policy-related decisions (12,13). While data is limited on the impact of COVID-19 with regards to barriers to working, it is indicated that healthcare providers are likely to suffer from high levels of anxiety and post-traumatic stress disorders (14).

Prevention remains the mainstay in the treatment and containment of the pandemic, requiring people at large to practice COVID-19 mitigating behaviors. Significant efforts hence need to be undertaken to strengthen beliefs about the disease in the population including the severity and susceptibility of threat so that people are more likely to take the needed actions to reduce the damage caused by the disease. As a result, it becomes important to study these key indicators to evaluate the public sense of threat to health, leading to the development of strategies for the general population and medical staff during COVID-19.

As psychological morbidity including depression, perceived stress and anxiety can compromise the social, emotional, psychological and physical functioning of a human being, the healthcare community is thought to be especially vulnerable. Psychological morbidity especially in the healthcare community may compromise patient care. It is therefore essential to carefully gauge the level of perceived susceptibility, severity, anxiety and subsequent response to COVID-19 between HCWs and non-healthcare workers (NHCWs). Identifying vulnerable areas and populations for psychological distress will help in strengthening health service delivery with targeted interventions. In this study we describe perceived severity, susceptibility and anxiety levels of HCWS in comparison to NHCWs. Furthermore, we explore vulnerable sub groups in the healthcare population with regards to training status, age, gender, income levels and level of patient care.

## Materials and Methods

### Study Design

A cross-sectional online survey was carried out in May 2020 through the social media channels of Aga Khan University (AKU), Pakistan, which included Facebook, Twitter, and LinkedIn. The survey link was also reposted on the Facebook page of Aga Khan University Hospital (AKUH), Pakistan.

### Study Participants

A convenience sampling strategy was used to enroll participants in the study. The online survey link remained active for two weeks. A total of 1405 respondents (507 HCWs and 899 NHCWs) filled the questionnaires. People aged 18 or above, residing in Pakistan for at least five days a week over the last month, with access to the internet and willing to participate in the survey were included in the study. The questionnaire was supposed to be filled once by each participant. Participants who could not respond to the study tool in either English or Urdu and participants who reported having filled the questionnaire at least once before were excluded from the study.

Respondents were categorized into NHCWs and HCWs. Those without basic (Bachelor level) training in any health or allied field were categorized as NHCWs whereas respondents having a formal training (students or graduates) in Medicine, Nursing, Pharmacy, Dentistry, Physiotherapy, Laboratory Technology or Allied Health Sciences including but not limited to homeopathy and *Hikmat* (a system of alternative medicine that involves the use of herbal remedies, dietary practices, and alternative therapies and addresses the prevention and treatment of disease) were categorized as HCWs. HCWs were further categorized into front-line and back-end HCWs. Front-line HCWs included all those professionals who are involved in direct patient care. Back-end HCWs included those who are currently not involved in direct clinical patient care including but not limited to undergraduate students of Medicine and Nursing and HCWs employed in the fields of Pharmacy, Dentistry, Physiotherapy, Laboratory Technology, Allied Health Sciences or others.

### Data Collection

Data was collected through two online self-administered semi-structured questionnaires (Questionnaire A and B). Questionnaire A was for NHCWs and Questionnaire B was for HCWs. Both questionnaires were developed on Google Forms. The questionnaires were adapted from the survey tool used in a similar study conducted in Hong Kong (21). Questionnaires A and B contained mostly identical questions, with a few additional questions in Questionnaire B related to HCWs’ field of study, training, place of work, perception about governmental measures, severity of COVID-19 as compared to other diseases and if they had acquired specific training related to COVID-19 in their respective organizations. Respondents, were asked about their demographics including gender, age, level of education, household income, permanent city of residence, and travel history followed by their health status in the past 14 days and whether they experienced any symptoms of illness.

Next, they were asked to rate the severity of the symptoms caused by COVID-19 and their perceived chance of survival if infected with COVID-19. Responses were captured using a five-point Likert scale. Subsequently, information exposure was probed by asking respondents about the sources from which they obtain information about COVID-19, and how reliable they deemed those sources to be. This was followed by questions on how likely one considered themselves and their families to be infected with COVID-19 if no preventive measures were taken. Participants’ anxiety and depression levels were assessed using the validated Hospital Anxiety and Depression Scale (HADS). This scale was used in a study conducted in Hong Kong for assessing anxiety related to COVID-19 (15,16). The possible minimum score for anxiety and depression is 0 and the maximum is 21. A score of 8 or above indicates anxiety. Respondents were also asked about the psychological impact of COVID-19 on their job, personal life, sleep, and eating habits.

### Data Analysis

Data collected from respondents was directly stored in Google Spreadsheets and later imported to Microsoft Excel and Statistical Package for the Social Sciences (SPSS) Version 21 (IBM Corp). Data was cleaned, coded, and analyzed using SPSS. A descriptive analysis was performed and results were tabulated as numbers (percentages) for qualitative variables and mean (±standard deviation) for quantitative variables. The Independent t-test or Mann Whitney U test or Pearson Chi-square test was applied to assess the differences between the groups’ (healthcare vs. NHCWs, healthcare students vs. graduates, frontline vs. back-end HCWs) perception of susceptibility and severity towards COVID-19, anxiety and depression level, the psychological impact of COVID-19, adoption of precautionary measures, reliability of information sources, and satisfaction with government measures. Bivariate and multivariate binary logistic regression analyses were performed to identify predictors (age, gender, household income, and presence of symptoms) of anxiety and depression among HCWs and NHCWs. Initially, in bivariate analysis, a single predictor at a time was entered and crude odds ratio (OR) and 95% confidence intervals (CI) were subsequently computed. A multivariate analysis, with all predictors entered at the same time, was completed to adjust for the effect of confounding, and adjusted OR and 95% CI were computed. All statistical tests were two-sided and a p-value of ≤0.05 was considered to be statistically significant.

### Ethical Consideration

Ethical approval was obtained from the Ethical Review Committee (ERC) of the Aga Khan University, Pakistan. Participants were asked to present their consent at the beginning of the survey and were free to withdraw at any stage.

## Results

Data from 1406 respondents (507 HCWs and 899 NHCWs) was analyzed. Majority of the respondents were males (53% HCWs and 72% NHCWs), below the age of 35 years (78% HCWs and 61% NHCWs), were residents of Karachi (49% HCWs and 50% NHCWs), and had a household income of ≤ PKR 40,000 (22% HCWs and 27% NHCWs) (Refer to Table 1). More than half of the HCWs (54%) belonged to the field of Medicine and 36% were currently working in a hospital, ward or a clinic (Refer to Table 2).

**Table 1.** Socio-demographic Characteristics of the Respondents.

| **Characteristics** | **HCWs**<br>**n = 507** | **NHCWs**<br>**n = 899** |
| --- | --- | --- |
|  | No. (%) | No. (%) |
| **Gender** |  |  |
| Male | 269 (53.1) | 644 (71.7) |
| Female | 235 (46.3) | 243 (27.0) |
| Prefer not to disclose | 3 (0.6) | 12 (1.3) |
| **Age** |  |  |
| 18-24years | 220 (43.4) | 239 (26.6) |
| 25-34 years | 173 (34.1) | 309 (34.4) |
| 35-44 years | 69 (13.6) | 213 (23.7) |
| 45-54years | 31 (6.1) | 73 (8.1) |
| 55 or above | 14 (2.8) | 61 (6.8) |
| Prefer not to disclose | 0 | 4 (0.4) |
| **Education** |  |  |
| Up to Matric/O-Levels |  | 18 (2.0) |
| Intermediate/A-Levels/International Baccalaureate |  | 110 (12.2) |
| Post-intermediate: Diploma/Certificate |  | 22 (2.5) |
| Bachelor or above |  | 743 (82.6) |
| Prefer not to disclose |  | 6 (0.7) |
| **Household incomes** |  |  |
| PKR 40,000 or below | 73 (22.4) | 246 (27.4) |
| PKR 40,001 – PKR 80,000 | 70 (21.4) | 176 (19.6) |
| PKR 80,001 – PKR 120,000 | 50 (15.4) | 156 (17.3) |
| ≥ PKR 120,001 | 56 (17.2) | 142 (15.8) |
| Prefer not to disclose | 77 (23.6) | 179 (19.9) |
| **Permanent residence** |  |  |
| a. Karachi | **249 (49.1)** | **451 (50.2)** |
| b. Lahore | 46 (9.0) | 82 (9.0) |
| c. Islamabad | 24 (4.8) | 61 (6.8) |
| d. Peshawar | 30 (5.9) | 23 (2.6) |
| e. Quetta | 8 (1.6) | 7 (0.8) |
| f. Hyderabad | 21 (4.2) | 32 (3.6) |
| g. Others | 129 (25.4) | 243 (27.0) |

**Table 2.** HCWs’ field, training status, and current work status (n=507)

| **Characteristics** | **No. (%)** |
| --- | --- |
| **Healthcare field** |  |
| Medicine | 274 (54.0) |
| Nursing | 52 (10.3) |
| Pharmacy | 42 (8.3) |
| Dentistry | 21 (4.1) |
| Physiotherapy | 24 (4.7) |
| Laboratory Technology or Allied Health Sciences | 87 (17.2) |
| Others | 7 (1.4) |
| **Training status** |  |
| Student | 181 (35.7) |
| Graduate | 326 (64.3) |
| **Current work status** |  |
| Working in hospital/ ward/clinic | 117 (35.9) |
| Working from home (online/ telephone etc.) | 49 (15.0) |
| Working in office setting | 28 (8.6) |
| Unpaid leave | 28 (8.6) |
| Paid leave | 18 (5.5) |
| Not working | 86 (26.4) |

### Perceived severity and susceptibility for COVID-19

No significant difference was observed between HCWs and NHCWs’ perception of susceptibility and severity towards COVID-19. About three-fourths of the respondents perceived that they (75% HCWs and 71% NHCWs) and their families (77% HCWs and 71% NHCWs) might get sick if they do not take preventive measures. Similarly, several respondents considered the symptoms of COVID-19 (if infected) as serious (46% HCWs and 38% NHCWs). Furthermore, most respondents thought that one could survive a COVID-19 infection (HCWs 70% and NHCWs 66%). However, a statistically significant difference was seen between the healthcare students’ and graduates’ perception of susceptibility and severity towards COVID-19. Healthcare graduates perceived themselves (66% students vs. 80% graduates, p-value 0.011) and their family (67% students vs. 82% graduates, p-value 0.008) to be more susceptible to COVID-19 than the healthcare students. Similarly, compared to students, more graduates perceived the disease to be severe (p-value 0.040). A significant difference was also seen between frontline and back-end HCWs’ perception of susceptibility and severity towards COVID-19. Frontline HCWs perceived themselves (83% frontline vs. 70% back-end, p-value 0.003) and their family (84% frontline vs. 72% back-end, p-value 0.006) as being more susceptible to COVID-19 than back-end HCWs. However, compared to those on the frontline, more back-end HCWs perceived the disease to be severe (p-value 0.045) (Refer to Table 3).

**Table 3.** Perceived severity and susceptibility for COVID-19.

| Variable | Perception | HCWs<br>n=507<br>No. (%) | NHCWs<br>n =899<br>No. (%) | HCWs<br>Vs<br>NHCW<br>s<br>P-value* | Health<br>care<br>Studen<br>ts<br>n = 181<br>No.<br>(%) | Healthcar<br>e<br>Graduate<br>s<br>n = 326<br>No. (%) | Studen<br>ts<br>Vs<br>Gradu<br>ates<br>P-value* | Frontli<br>ne<br>HCWs<br>n=216<br>No. (%) | Backend<br>HCWs<br>n=290<br>No. (%) | Frontli<br>ne Vs<br>Backen<br>d<br>P-value* |
| --- | --- | --- | --- | --- | --- | --- | --- | --- | --- | --- |
| Susceptibility |  |  |  |  |  |  |  |  |  |  |
| 1. I might contract the disease if no preventive measure is taken | Agree | 382 (75.3) | 636 (70.7) | 0.506 | 120 (66.3) | 262 (80.4) | 0.011 | 179 (82.9) | 203 (69.8) | 0.003 |
|  | Neutral | 41 (8.1) | 128 (14.3) |  | 18 (9.9) | 23 (7.1) |  | 17 (7.9) | 24 (8.2) |  |
|  | Disagree | 81 (16.0) | 129 (14.3) |  | 42 (23.2) | 39 (11.9) |  | 20 (9.2) | 61 (21.0) |  |
|  | Don't know | 3 (0.6) | 6 (0.7) |  | 1 (0.6) | 2 (0.6) |  | 0 | 3 (1.0) |  |
| 2. My family might contract the disease if no preventive measure is taken | Agree | 389 (76.7) | 642 (71.4) | 0.539 | 121 (66.8) | 268 (82.2) | 0.008 | 181 (83.8) | 208 (71.5) | 0.006 |
|  | Neutral | 31 (6.1) | 116 (12.9) |  | 15 (18.3) | 16 (4.9) |  | 10 (4.6) | 21 (7.2) |  |
|  | Disagree | 84 (16.6) | 133 (14.8) |  | 44 (24.3) | 40 (12.3) |  | 25 (11.6) | 59 (20.3) |  |
|  | Don't know | 3 (0.6) | 8 (0.9) |  | 1 (0.6) | 2 (0.6) |  | 0 | 3 (1.0) |  |
| 3. I might contract COVID-19 if one of my family members tests positive for the disease | Agree | 350 (69.3) | 594 (66.1) | 0.559 | 118 (65.6) | 232 (71.4) | 0.637 | 158 (73.5) | 192 (66.3) | 0.138 |
|  | Neutral | 60 (11.9) | 140 (15.5) |  | 18 (10.0) | 42 (12.9) |  | 24 (11.2) | 36 (12.4) |  |
|  | Disagree | 79 (15.6) | 139 (15.5) |  | 38 (21.1) | 41 (12.6) |  | 27 (12.5) | 52 (17.9) |  |
|  | Don't know | 16 (3.2) | 26 (2.9) |  | 6 (3.3) | 10 (3.1) |  | 6 (2.8) | 10 (3.4) |  |
| Severity |  |  |  |  |  |  |  |  |  |  |
| 1. Seriousness of symptoms caused by SARS-CoV 19 | Severe | 232 (45.8) | 342 (38.0) | 0.916 | 96 (53.0) | 136 (41.7) | 0.04 | 93 (43.1) | 139 (47.9) | 0.045 |
|  | Neutral | 148 (29.2) | 192 (21.4) |  | 45 (24.9) | 103 (31.6) |  | 64 (29.6) | 84 (28.9) |  |
|  | Not Severe | 82 (16.1) | 127 (14.1) |  | 23 (12.7) | 59 (18.1) |  | 44 (20.4) | 38 (13.0) |  |
|  | Don't know | 45 (8.9) | 238 (26.5) |  | 17 (9.4) | 28 (8.6) |  | 15 (6.9) | 30 (10.2) |  |
| 2. Chance of survival if infected with COVID-19 | High | 356 (70.2) | 596 (66.3) | 0.807 | 124 (68.5) | 232 (71.2) | 0.46 | 147 (68.1) | 209 (71.8) | 0.188 |
|  | Neutral | 105 (20.7) | 173 (19.2) |  | 40 (20.1) | 65 (19.9) |  | 49 (22.7) | 56 (19.3) |  |
|  | Not high | 34 (6.7) | 51 (5.7) |  | 13 (7.2) | 21 (6.4) |  | 17 (7.9) | 17 (5.8) |  |
|  | Don't know | 12 (2.4) | 79 (8.8) |  | 4 (2.2) | 8 (2.5) |  | 3 (1.3) | 9 (3.1) |  |

\*Mann-Whitney test
Note: Categories were merged, so “agree/severe/high” and “strongly agree/ very severe/ very high” were merged into category
“agree/severe/high”, and categories “disagree/ not severe/ not high” and “strongly disagree/ not severe at all/ not high at all” were
merged into “disagree/ not severe/ not high”.

### Psychological Distress in HCWs and NHCWs

Over half of the respondents were found to be either anxious (54% HCWs and 59% NHCWs) or depressed (54% HCWs and 57% NHCWs) as indicated by the HADS scores. No significant difference was seen in the anxiety and depression levels of HCWs and NHCWs. However, the incidence of depression was significantly higher among healthcare students compared to healthcare graduates (Mean (SD): 8.40 (3.45) in students 7.72 (3.80) in graduates, p-value 0.047). Around 62% of healthcare students and 49% of graduates had depression. A statistically significant difference was noted between frontline and back-end HCWs’ perception about the impact of COVID-19 on their personal life (75% frontline vs. 58% back-end HCWs, p-value <0.001). However, no significant difference was reported between HCWs’ and NHCWs’ perceived impact of COVID-19 on their jobs, personal life, sleeping pattern, and eating habits (Refer to Table 4).

**Table 4.**
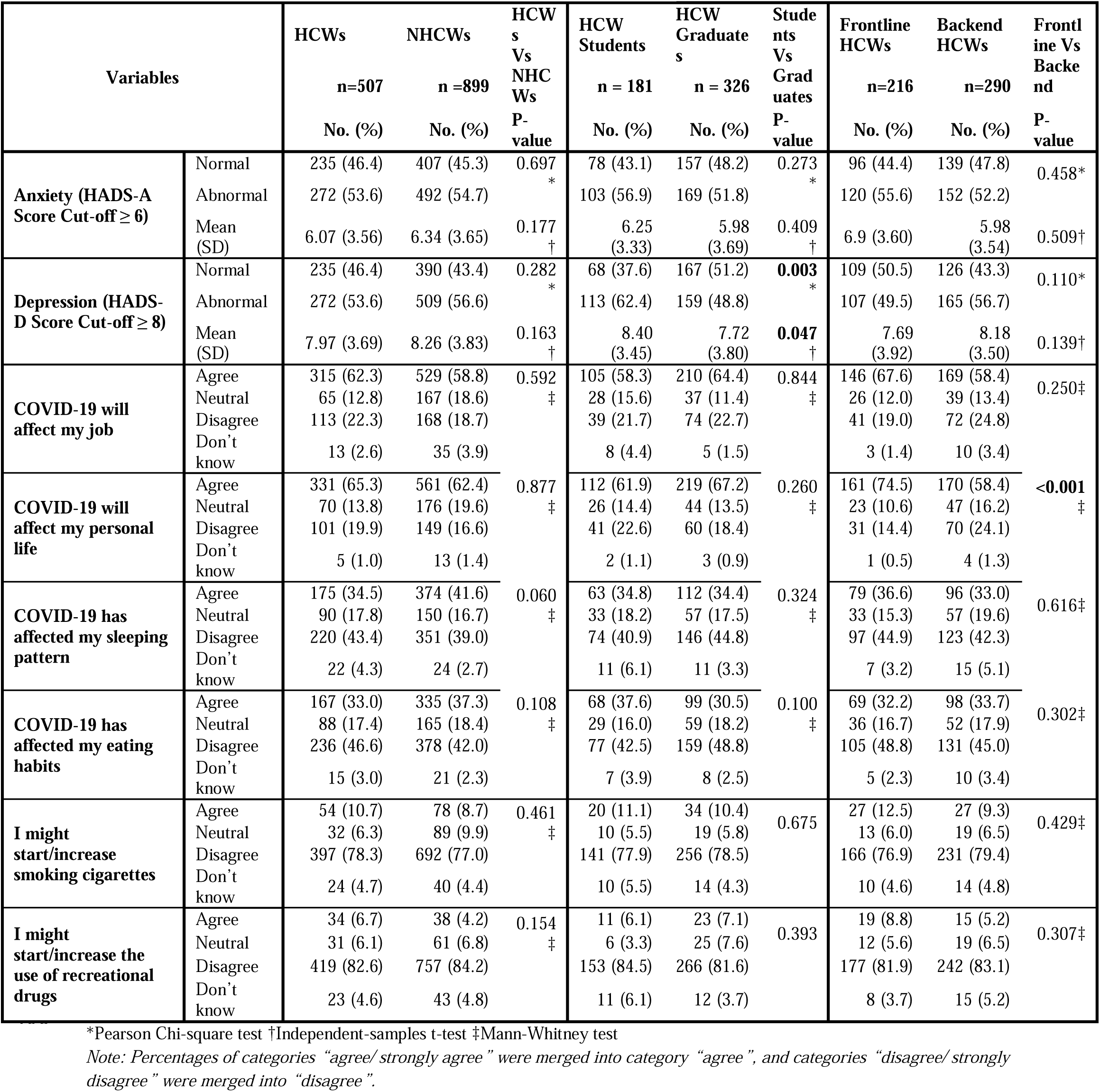
Perceived Psychological Impact of COVID-19.

### Predictors of Psychological Distress in HCWs and NHCWs

Gender, age, and presence of symptoms had significant positive associations with anxiety among HCWs. Female HCWs were nearly twice as likely to be anxious than the male HCWs (aOR: 2.34, 95% CI: 1.37-3.99, p-value 0.002). HCWs of younger age (25-34 years) (aOR 3.44, 95% CI: 1.30-9.09, p-value: 0.013) were nearly three times more likely to have anxiety than HCWs of 55 years or above. HCWs having COVID-19 related symptoms were 2.09 times more likely to have anxiety than HCWs without symptoms (aOR: 2.09, 95% CI: 1.01-4.32, p-value: 0.046) (Refer to table 5).

**Table 5.** Predictors of Anxiety and Depression.

| Variable | Anxiety |  |  |  | Depression |  |  |  |
| --- | --- | --- | --- | --- | --- | --- | --- | --- |
|  | HCWs<br>(n=507) |  | NHCWs<br>(n=899) |  | HCWs |  | NHCWs |  |
|  | aOR* (95%<br>CI) | P-value | aOR* (95%<br>CI) | P-value | aOR* (95%<br>CI) | P-value | aOR* (95%<br>CI) | P-value |
| **Gender** |  |  |  |  |  |  |  |  |
| Female | 2.34 (1.37-3.99) | **0.002** | 1.62 (1.12-2.35) | **0.010** | 1.53 (0.90-2.58) | 0.115 | 1.41 (0.98-2.02) | 0.065 |
| Male | Reference |  | Reference |  | Reference |  | Reference |  |
| **Age** |  |  |  |  |  |  |  |  |
| 18-24years | 3.52 (1.19-10.42) | **0.023** | 2.53 (1.51-4.23) | **<0.001** | 1.00 (0.37-2.71) | 1.000 | 1.38 (0.84-2.27) | 0.209 |
| 25-34 years | 3.44 (1.30-9.09) | **0.013** | 2.84 (1.75-4.62) | **<0.001** | 1.72 (0.72-4.11) | 0.219 | 1.26 (0.79-2.01) | 0.325 |
| 35-44 years | 4.68 (1.63-13.47) | **0.004** | 2.21 (1.31-3.70) | **0.003** | 1.01 (0.39-2.64) | 0.978 | 1.66 (1.00-2.76) | **0.050** |
| 45 years or above | Reference |  | Reference |  | Reference |  | Reference |  |
| **Household incomes** |  |  |  |  |  |  |  |  |
| ≤ PKR 60,000 | 1.30 (0.62-2.71) | 0.491 | 1.61 (1.06-2.43) | **0.024** | 1.12 (0.54-2.32) | 0.762 | 1.58 (1.06-2.36) | **0.026** |
| PKR 60,001 – PKR 120,000 | 1.35 (0.64-2.84) | 0.430 | 2.22 (1.42-3.48) | **<0.001** | 1.25 (0.61-2.57) | 0.548 | 2.29 (1.48-3.54) | **<0.001** |
| > PKR 120,000 | Reference |  | Reference |  | Reference |  | Reference |  |
| **Presence of COVID-19 related Symptoms** |  |  |  |  |  |  |  |  |
| Yes | 2.09 (1.01-4.32) | **0.046** | 1.98 (1.34-2.94) | **0.001** | 2.72 (1.34-5.55) | **0.006** | 1.41 (0.96-2.06) | 0.077 |
| No | Reference |  | Reference |  | Reference |  | Reference |  |

\*Adjusted for gender, age, household income, and presence of COVID-19 related symptoms.

Similarly, gender, age, household income, and presence of symptoms were positively associated with anxiety among NHCWs. Female NHCWs were 1.62 times more likely to have anxiety than male NHCWs (aOR: 1.62, 95% CI: 1.12-2.35, p-value 0.010). NHCWs of younger age (25-34 years) (aOR 2.84, 95% CI: 1.75-4.62, p-value: <0.001) were nearly three times more likely to be anxious than NHCWs of 55 years or above. NHCWs having income level of 60,001-120,000 PKR were 2.22 times more likely to have anxiety than NHCWs having household income ≥ PKR 120,000 PKR (aOR: 2.22, 95% CI: 1.42-3.48, p-value: <0.001). NHCWs having COVID-19 related symptoms were 1.98 times more likely to have anxiety than HCWs without symptoms (aOR: 1.98; 95% CI: 1.34-2.94, p-value: 0.001) (Refer to table 5).

Furthermore, presence of symptoms was positively associated with depression among HCWs (aOR: 2.72; 95% CI: 1.34-5.55, p-value: 0.006). Household income had a positive association with depression among NHCWs. NHCWs having income level of 60,001-120,000 PKR were nearly two times more likely to have depression than NHCWs having household income > PKR 120,000 PKR (aOR: 2.29, 95% CI: 1.48-3.54, p-value: <0.001) (Refer to table 5).

### Adoption of Precautionary Measures

Significantly more HCWs reported wearing face masks (94% HCWs vs. 91% NHCWs, p-value 0.012), avoiding visiting meat shops or markets (77% HCWs vs. 66% NHCWs, p-value <0.001) than NHCWs. Moreover, significantly less HCWs reported that they refrain from going to hospitals or clinics (60% HCWs vs. 81% NHCWs, p-value <0.001) and work (55% HCWs vs. 66% NHCWs, p-value<0.001) compared to NHCWs. Additionally, there was a statistically significant difference between healthcare students’ and graduates’ adoption of some precautionary measures such as washing their hands with soap/sanitizer frequently (96% students vs. 99% graduates, p-value 0.001), avoiding going out (87% students vs. 73% graduates, p-value 0.003), refraining from going to hospital or clinic (80% students vs. 50% graduates, p-value <0.001) Similarly, a statistically significant difference was noted between frontline and back-end HCWs in the adoption of some precautionary measures such as washing their hands with soap/sanitizer frequently (100% frontline vs. 97% back-end, p-value 0.009, refraining from going to hospital or clinic (45% frontline vs. 72% back-end, p-value<0.001) and avoiding going to work (37% frontline vs. 68% back-end, p-value<0.00)1 (Refer to Table 6).

**Table 6.** Adoption of Precautions by the respondents (Number of respondents answering Yes”)

| Precautions | HCWs<br>n=507<br>No. (%) | NHCWs<br>n =899<br>No. (%) | HCWs<br>Vs<br>NHCWs<br>P-value* | Healthcare<br>Students<br>n = 181<br>No. (%) | Healthcare<br>Graduates<br>n = 326<br>No. (%) | Students<br>Vs<br>Graduates<br>P-value* | Frontline<br>NHCWs<br>n=216<br>No. (%) | Backend<br>NHCWs<br>n=290<br>No. (%) | Frontline<br>Vs<br>Backend<br>P-value* |
| --- | --- | --- | --- | --- | --- | --- | --- | --- | --- |
| 1. Wear face masks | 475 (93.7) | 815 (90.7) | **0.012** | 161<br>(89.0) | 314 (96.3) | 0.082 | 208 (96.3) | 267<br>(91.8) | 0.344 |
| 2. Wash hands frequently<br>(With soap or hand<br>sanitizer) | 497 (98.0) | 884 (98.3) | 0.756 | 173<br>(95.6) | 324 (99.4) | **0.001** | 215 (99.5) | 282<br>(96.9) | **0.009** |
| 3. Avoid contacting people<br>who have fever or<br>respiratory symptoms | 470 (92.7) | 825 (91.8) | 0.597 | 163<br>(90.1) | 307 (94.2) | **0.047** | 203 (94.0) | 267<br>(91.8) | 0.212 |
| 4. Avoid going out | 397 (78.3) | 679 (75.5) | 0.089 | 158<br>(87.3) | 239 (73.3) | **0.003** | 164 (75.9) | 233<br>(80.1) | 0.685 |
| 5. Avoid going to meat<br>shops/market | 391 (77.1) | 592 (65.9) | **<0.001** | 142<br>(78.5) | 249 (76.4) | 0.291 | 172 (79.6) | 219<br>(75.3) | 0.330 |
| 6. Avoid going to hospital<br>or clinic | 306 (60.4) | 728 (81.0) | **<0.001** | 144<br>(79.6) | 162 (49.7) | **<0.001** | 97 (44.9) | 209<br>(71.8) | **<0.001** |
| 7. Avoid taking public<br>transportation | 456 (89.9) | 838 (93.2) | **<0.021** | 165<br>(91.2) | 291 (89.3) | 0.651 | 193 (89.4) | 263<br>(90.4) | 0.207 |
| 8. Avoid going to work | 277 (54.6) | 594 (66.1) | **<0.001** | 137<br>(75.7) | 140 (42.9) | **<0.001** | 80 (37.0) | 197<br>(67.7) | **<0.001** |
| 9. Avoid going to school or<br>avoid letting children go<br>to school | 382 (75.3) | 811 (90.2) | **0.001** | 154<br>(85.1) | 228 (69.9) | **0.024** | 149 (69.0) | 233<br>(80.1) | 0.733 |
| 10. Avoid international travel | 467 (92.1) | 853 (94.9) | **0.032** | 169<br>(93.4) | 298 (91.4) | 0.954 | 198 (91.7) | 269<br>(92.4) | 0.980 |
| 11. Avoid domestic or inter-<br>city travel | 440 (86.8) | 805 (89.5) | 0.076 | 165<br>(91.2) | 275 (84.4) | 0.091 | 174 (80.6) | 266<br>(91.4) | **0.001** |

\* Pearson Chi-square test
Note: Only the most salient variables for social distancing have been reported in this table.

### Sources of Information utilized by the Respondents

Nearly all HCWs and NHCWs remained alert to the disease progression of COVID-19 (96% for the former and 95% of the latter). The most trusted sources of information were doctors (91% HCWs and 87% NHCWs, p-value 0.003) and government websites (85% HCWs and 80% NHCWs, p-value 0.056). Furthermore, significantly more NHCWs considered magazine (39% HCWs vs. 44% NHCWs, p value 0.005), television (54% HCWs vs. 63% NHCWs, p value 0.043), family or friends (46% HCWs vs. 54% NHCWs, p value 0.001) as reliable sources of information. Likewise, a statistically significant percentage of healthcare students considered official government websites (86% students vs. 84% graduates), television (60% students vs. 51% graduates, p-value 0.040), radio (58% students vs. 44% graduates, p-value; 0.017), and magazine (50% students vs. 33% graduates, p-value; 0.015) as reliable sources of information compared to healthcare graduates. Moreover, compared to back-end HCWs, more frontline HCWs believed in the reliability of information received through television (47% frontline vs. 59% back-end, p-value 0.049), radio (42% frontline vs. 54% back-end, p-value 0.046), newspaper (49% frontline vs. 62% back-end, p-value 0.015), and magazine (28% frontline vs. 47% back-end, p-value 0.015) (Refer to Table 7).

**Table 7.** Perceived reliability of information sources.

| Information Source | Perception | HCWs<br>n=488<br>No. (%) | NHCWs<br>n =864<br>No. (%) | HCWs<br>Vs<br>NHCW<br>s<br>P-value* | Healthcar<br>e<br>Students<br>n = 175<br>No. (%) | Healthcar<br>e<br>Graduate<br>s<br>n = 313<br>No. (%) | Student<br>s<br>Vs<br>Gradua<br>tes<br>P-value* | Frontline<br>HCWs<br>n=206<br>No. (%) | Backend<br>HCWs<br>n=282<br>No. (%) | Fr<br>ne<br>Vs<br>Ba<br>d<br>v |
| --- | --- | --- | --- | --- | --- | --- | --- | --- | --- | --- |
| Newspaper | Reliable | 268 (56.5) | 500 (57.9) | 0.239 | 105 (63.2) | 163 (52.9) | 0.071 | 100 (49.0) | 168 (62.2) |  |
|  | Neutral | 138 (29.2) | 273 (31.7) |  | 36 (21.7) | 102 (33.1) |  | 77 (37.8) | 61 (22.6) |  |
|  | Unreliable | 68 (14.3) | 90 (10.4) |  | 25 (15.1) | 43 (14.0) |  | 27 (13.2) | 41 (15.2) |  |
| Magazine | Reliable | 189 (39.0) | 378 (43.7) | 0.005 | 87 (50.3) | 102 (32.7) | 0.015 | 58 (28.3) | 131 (46.8) |  |
|  | Neutral | 176 (36.3) | 347 (40.2) |  | 44 (25.4) | 132 (42.3) |  | 94 (45.8) | 82 (29.3) |  |
|  | Unreliable | 120 (24.7) | 139 (16.1) |  | 42 (24.3) | 78 (25.0) |  | 53 (25.9) | 67 (23.9) |  |
| Radio | Reliable | 238 (49.0) | 463 (53.6) | 0.051 | 100 (57.5) | 138 (44.2) | 0.017 | 86 (42.0) | 152 (54.1) |  |
|  | Neutral | 163 (33.5) | 311 (36.0) |  | 44 (25.3) | 119 (38.1) |  | 84 (41.0) | 79 (28.1) |  |
|  | Unreliable | 85 (17.5) | 90 (10.4) |  | 30 (17.2) | 55 (17.6) |  | 35 (17.0) | 50 (17.8) |  |
| Television | Reliable | 262 (53.9) | 546 (63.2) | 0.043 | 104 (59.8) | 158 (50.6) | 0.040 | 97 (47.3) | 165 (58.7) |  |
|  | Neutral | 141 (29.0) | 186 (21.5) |  | 43 (24.7) | 98 (31.4) |  | 73 (35.6) | 68 (24.2) |  |
|  | Unreliable | 83 (17.1) | 132 (15.3) |  | 27 (15.5) | 56 (18.0) |  | 35 (17.1) | 48 (17.1) |  |
| Government websites | Reliable | 411 (84.6) | 693 (80.2) | 0.056 | 150 (86.2) | 261 (83.7) | 0.001 | 168 (81.9) | 243 (86.5) |  |
|  | Neutral | 48 (9.9) | 110 (12.7) |  | 13 (7.5) | 35 (11.2) |  | 25 (12.2) | 23 (8.2) |  |
|  | Unreliable | 27 (5.5) | 61 (7.1) |  | 11 (6.3) | 16 (5.1) |  | 12 (5.9) | 15 (5.3) |  |
| Unofficial websites | Reliable | 134 (27.7) | 237 (27.4) | 0.156 | 59 (33.9) | 75 (24.2) | 0.983 | 41 (20.2) | 93 (33.1) |  |
|  | Neutral | 118 (24.4) | 282 (32.7) |  | 28 (16.1) | 90 (29.0) |  | 61 (30.0) | 57 (20.3) |  |
|  | Unreliable | 232 (47.9) | 345 (39.9) |  | 87 (50.0) | 145 (46.8) |  | 101 (49.8) | 131 (46.6) |  |
| Social media platforms | Reliable | 149 (30.7) | 273 (31.6) | 0.004 | 65 (37.1) | 84 (27.0) | 0.358 | 50 (24.5) | 99 (35.1) |  |
|  | Neutral | 96 (19.7) | 276 (31.9) |  | 23 (13.2) | 73 (23.5) |  | 54 (26.5) | 42 (14.9) |  |
|  | Unreliable | 241 (49.6) | 315 (36.5) |  | 87 (49.7) | 154 (49.5) |  | 100 (49.0) | 141 (50.0) |  |
| Doctor | Reliable | 443 (91.2) | 753 (87.2) | 0.003 | 158 (90.3) | 285 (91.6) | 0.942 | 186 (90.7) | 257 (91.4) |  |
|  | Neutral | 35 (7.2) | 97 (11.2) |  | 12 (6.9) | 23 (7.4) |  | 16 (7.8) | 19 (6.8) |  |
|  | Unreliable | 8 (1.6) | 14 (1.6) |  | 5 (2.8) | 3 (1.0) |  | 3 (1.5) | 5 (1.8) |  |
| Family or friends | Reliable | 221 (45.7) | 464 (53.8) | 0.001 | 74 (42.3) | 147 (47.6) | 0.185 | 85 (41.9) | 136 (48.4) |  |
|  | Neutral | 158 (32.6) | 290 (33.6) |  | 57 (32.6) | 101 (32.7) |  | 71 (34.9) | 87 (31.0) |  |
|  | Unreliable | 105 (21.7) | 109 (12.6) |  | 44 (25.1) | 61 (19.7) |  | 47 (23.2) | 58 (20.6) |  |

\*Mann Whitney test
Note: Percentages of categories “reliable/ very reliable” were merged into category “reliable”, and categories “unreliable / very unreliable” were merged into “unreliable”.

### Satisfaction with Government Measures

Less than a third of HCWs and NHCWs were satisfied with the government’s measures to control COVID-19. HCWs, in comparison with NHCWs, were significantly more dissatisfied with the availability of Personal Protective Equipment (62% vs. 46%, p value <0.001), testing kits (49% vs. 41%, p value 0.028), and screening facilities (54% vs. 42%, p value <0.001). Similarly, compared to healthcare students, graduates were significantly more dissatisfied with screening facilities (57% graduates vs. 49% students, p-value0.016), testing kits (52% graduates vs. 43% students, p-value 0.016), and quarantine facilities (49% graduates vs. 38% students, p-value 0.012). However, no significant difference was seen between frontline and back-end HCWs’ satisfaction with government’s measures to control COVID-19 (Refer to Table 8).

**Table 8.** Respondents’ satisfaction with government measures.

| Measures | HCWs<br>n=507<br>No. (%) | NHCWs<br>n =589<br>No. (%) | HCWs<br>Vs<br>NHCW<br>s<br>P-value* | Healthcar<br>e<br>Students<br>n = 181<br>No. (%) | Healthca<br>re<br>Graduate<br>s<br>n = 326<br>No. (%) | Student<br>s<br>Vs<br>Gradua<br>tes<br>P-value* | Frontline<br>NHCWs<br>n=216<br>No. (%) | Backend<br>NHCWs<br>n=290<br>No. (%) | Frontli<br>ne<br>Vs<br>Backen<br>d<br>P-value* |
| --- | --- | --- | --- | --- | --- | --- | --- | --- | --- |
| **Screening facilities** |  |  |  |  |  |  |  |  |  |
| Satisfied/ Very Satisfied | 104 (20.6) | 111 (18.8) | <0.001 | 46 (25.6) | 58 (17.9) | **0.016** | 39 (18.2) | 65 (22.4) | 0.245 |
| Neutral | 117 (23.2) | 198 (33.6) |  | 44 (24.4) | 73 (22.5) |  | 49 (23.0) | 68 (23.4) |  |
| Unsatisfied/ Very Unsatisfied | 273 (54.2) | 248 (42.2) |  | 89 (49.4) | 184 (56.8) |  | 124 (57.9) | 149 (51.4) |  |
| Don't know | 10 (2.0) | 32 (5.4) |  | 1 (0.6) | 9 (2.8) |  | 2 (0.9) | 8 (2.8) |  |
| **Laboratory services/ testing kits** |  |  |  |  |  |  |  |  |  |
| Satisfied/Very Satisfied | 109 (21.6) | 124 (21.0) | **0.028** | 50 (27.8) | 59 (18.1) | **0.016** | 41 (19.1) | 68 (23.4) | 0.344 |
| Neutral | 134 (26.5) | 191 (32.4) |  | 48 (26.7) | 86 (26.5) |  | 60 (27.9) | 74 (25.6) |  |
| Unsatisfied/ Very Unsatisfied | 248 (49.1) | 243 (41.3) |  | 78 (43.3) | 170 (52.3) |  | 111 (51.6) | 137 (47.2) |  |
| Don't know | 14 (2.8) | 31 (5.3) |  | 4 (2.2) | 10 (3.1) |  | 3 (1.4) | 11 (3.8) |  |
| **Quarantine facilities** |  |  |  |  |  |  |  |  |  |
| Satisfied/Very Satisfied | 134 (26.6) | 157 (26.6) | **0.022** | 56 (31.1) | 78 (24.1) | **0.012** | 56 (26.2) | 78 (26.9) | 0.602 |
| Neutral | 125 (24.8) | 179 (30.4) |  | 50 (27.8) | 75 (23.1) |  | 51 (23.8) | 74 (25.5) |  |
| Unsatisfied/ Very Unsatisfied | 228 (45.2) | 223 (37.9) |  | 69 (38.3) | 159 (49.1) |  | 103 (48.1) | 125 (43.1) |  |
| Don't know | 17 (3.4) | 30 (5.1) |  | 5 (2.8) | 12 (3.7) |  | 4 (1.9) | 13 (4.5) |  |
| **Personal Protective Equipment** |  |  |  |  |  |  |  |  |  |
| Satisfied/Very Satisfied | 86 (17.0) | 100 (17.0) | <0.001 | 39 (21.5) | 47 (14.5) | 0.072 | 29 (13.6) | 57 (19.6) | 0.076 |
| Neutral | 95 (18.8) | 186 (31.6) |  | 38 (21.0) | 57 (17.6) |  | 37 (17.3) | 58 (19.9) |  |
| Unsatisfied/ Very Unsatisfied | 311 (61.6) | 268 (45.5) |  | 102 (56.4) | 209 (64.5) |  | 144 (67.3) | 167 (57.4) |  |
| Don't know | 13 (2.6) | 35 (5.9) |  | 2 (1.1) | 11 (3.4) |  | 4 (1.8) | 9 (3.1) |  |

\*Mann-Whitney test † Pearson Chi-square test
Note: Only the most salient variables have been reported in this table. Percentages of categories "satisfied" and "very satisfied" were
merged into category "satisfied /very satisfied", and categories "unsatisfied" and "very unsatisfied" were merged into
"unsatisfied/very unsatisfied".

## Discussion

The COVID-19 pandemic has caused a global health crisis. As the disease burden exponentially increases, HCWs find themselves susceptible to contracting the disease or being the source of exposure for their family members. This constant threat puts them at a high risk of psychological distress. This study explores the similarities and differences in risk perceptions, anxiety levels, and behavioral responses of HCWs and NHCWs in Pakistan during the COVID-19 pandemic. Respondents’ adoption of precautionary measures and satisfaction with the government’s response were also assessed.

This study showed that the perceived susceptibility and severity towards COVID-19 was high in both HCWs and NHCWs. Healthcare graduates perceived themselves and their families to be more susceptible to COVID-19 and found the disease to be much more severe than healthcare students. A study from Hong Kong showed an even higher perception of susceptibility and severity towards COVID-19 (15,16). The generally high-risk perception levels can be explained by the coverage of the pandemic on social media as well as television sensitizing the people to the disease. Frequency of watching the media and sources of information has been known to influence risk perception (17). This is supported by reports from the MERS, SARS and H1N1 outbreaks (18,19).

Additionally, frontline HCWs perceived themselves and their families to be more susceptible to COVID-19 than back-end HCWs while the latter perceived the disease to be more severe. Similar to HCWs in this study, training status and clinical placement creates differences in risk perception, as seen in the medical students of Iran (20). This indicates that HCWs felt more vulnerable with greater exposure to the infected individuals. Direct contact with COVID-19 patients is hence a major cause for concern among HCWs for themselves and their families. Greater perceived severity among backend workers on the other hand may be explained by the fact that since these workers are not seeing patients recover as frequently, their notion of disease severity is higher.

More than half the respondents in the study had some form of psychological distress (anxiety or depression). Data describing gaps in anxiety levels between HCWs and NHCWs is limited. Contrary to this study, which showed that both HCWs and NHCWs had similar high anxiety levels, HCWs in Italy reported higher anxiety levels in comparison to the general population (21). Therefore, the gap in anxiety levels seen in Italy (between HCWs and general population) is not seen in Pakistan. This Pakistan reported a lower disease burden compared to Italy, therefore it is possible that the HCWs in Pakistan are generally less anxious due to a lesser case load and severity at the time of data collection. A possible reason for higher anxiety levels in the NHCWs group in this study could be due to higher sensitization by the social media creating undue anxiety. Another possible reason for higher anxiety levels in the NHCWs group in this study could be a higher average education level (Bachelor’s) which is an independent risk factor for elevated anxiety symptoms (22).

Both HCWs and NHCWs had similar perceptions of the impact of COVID-19 on their daily routine. The perceived impact, however, was greater among frontline HCWs compared to the back-end HCWs. Frontline HCWs (doctors and nurses) are involved in more direct patient care and have greater patient interaction. New protocols and added personal protective equipment (PPE) are focused on frontline workers more which warrants a greater transition from the pre-pandemic life. While backend HCWs (pharmacists, dentists, physiotherapists, allied health sciences, and students) also had additions in their daily routine such as masks, social distancing, and hand sanitizing, these changes are comparatively less cumbersome compared to changes in frontline HCW’s routines. The greater the patient interaction, greater number of precautions are required in daily routine which results in greater impact on personal lives. A study reported nurses having more anxiety in comparison to doctors as they are involved for a longer duration with the patients (23). Additionally, frontline HCWs perceived themselves and their families to be more susceptible to COVID-19 than back-end HCWs while the latter perceived the disease to be more severe. Greater perceived severity among backend workers on the other hand may be explained by the fact that since these workers are not seeing patients recover as frequently, their notion of disease severity is higher.

This study pointed out that healthcare students were more depressed than the graduates. Existing data that has generally shown students as a high risk group for depression (24). Experiences from the past epidemics also provide similar evidence (25–27). All educational institutes in Pakistan were closed during the duration of the study. Although educational institutes adapted to online classes and virtual examinations, it took a long while before students could get used to new routines and methods. These interruptions in schedules, lack of physical interaction with peers and social isolation may have contributed to the greater depression levels. Furthermore, students are often more active on social media which has been a great source of information and sensitization of COVID-19 pandemic. Constant ill news and disturbing figures may have further added to the current depression levels of healthcare students. Healthcare students are required to rotate in pre-scheduled clinical clerkships which mandates interactive patient care. Completing clinical clerkships require patient interaction and are practically impossible in an online virtual setting which have led to delayed graduations. This uncertainty and implications for a wasted academic year may also contribute to the increased depression levels in healthcare students.

Female gender, younger age, and presence of COVID-19 related symptoms predicted increased psychological distress in HCWs while lower-income and presence of COVID-19 related symptoms predicted the same in NHCWs. Female gender has also been linked with greater anxiety levels in Iran and China (24). Unlike our results, age did not predict psychological distress in the Chinese population. The differences in the predictors of distress during the COVID-19 pandemic could be attributed to the differences in countries’ healthcare systems, the availability of personal protective equipment (PPE), cultures, employment conditions, lockdown and work from home policies, maintaining a living in a pandemic and mainstream and social media information, to name just a few. COVID-19 has also increased the financial burden on households as many people struggle to run small businesses and maintain daily income. The fear of not being able to fulfil the basic necessities may be the reason why lower income can increase anxiety levels. The results, therefore, suggest the need for the identification of useful predictors of mental health in individual countries during the COVID-19 pandemic.

Only less than a third of HCWs and NHCWs were satisfied with the government’s measures to control COVID-19. HCWs, in comparison with NHCWs, were significantly more dissatisfied with the availability of PPEs and screening facilities. Pakistan is a resource-limited country with no prior experience of handling a pandemic which may explain the lack of satisfaction with the government’s response. This is concerning because healthcare staff’s access to PPEs predicts lower distress levels, better physical health conditions, and more job satisfaction (28). Moreover, frontline HCWs who reuse or have inadequate access to PPE are known to have the higher risk of COVID-19 infection (29). Therefore, it is important for the government to address the concerns particularly among HCWs as they are the foot soldiers fighting the pandemic.

The main strength of this study is the comparison of various sub-groups within HCWs for a comprehensive analysis. In this respect, it is a novel study accounting for differences in experiences among HCWs. However, frontline and back-end categorization was made without using any standard classification. Moreover, in this study, most of the respondents were aged less than 35 years, which may not accurately represent the older population who are at greater risk for contracting COVID-19. Nevertheless, as the majority of the population in Pakistan is below the age of 30 years, respondents are likely to represent the perceptions of the literate people. Lastly, this was an online survey with most respondents either having a Bachelor’s degree or above. This may result in inadequately justifying the attitudes and perceptions of the NHCWs as the Pakistani population on average has a much lower educational background.

To decrease the level of psychological distress, hospital administrators should implement policies to target mental well-being of the HCWs. Hospital staff dealing with COVID-19 patients should be monitored on a regular basis to avoid burnout. Incentives such as financial bonuses and paid leaves should be provided. To cater for the individual needs of the population, communities should establish online mental health and wellness groups to overcome social isolation. Government should ensure provision of PPE, testing kits, and screening facilities to increase satisfaction levels of HCWs in particular and the public at large. Furthermore, implementing these strategies may also contribute in mitigating the disease spread. The better the disease is controlled, the lesser will be the psychological morbidity and adverse impact on people’s lives.

## Conclusion

HCWs and NHCWs both have high levels of perceived susceptibility and severity along with increased psychological distress. This study also identified vulnerable groups such as frontline HCWs, healthcare students, younger aged people, females, and individuals with lower income to be at a higher risk of psychological distress. We recommend exploring other vulnerable groups not evaluated in this study such as COVID-19 positive patients, older and immunocompromised patients for risk perceptions and psychological distress. Further studies need to investigate a direct link between HCWs and the development of COVID-19 infection to quantify the infection risk. HCWs are the frontline fighters against pandemics and preventing their psychological morbidity must be prioritized for ensuring their well-being as well as that of patients that they treat within the health care system.

## Supporting information

STROBE checklist for cross-sectional studies

## Data Availability

Data and materials are in the custody of the University and could be made available on request, keeping the individual identity of the participants confidential.

## Conflict of Interest

The authors declare that the research was conducted in the absence of any commercial or financial relationships that could be construed as a potential conflict of interest.

## Author Contributions

FR conceived the study, guided data collection and reviewed all drafts of the manuscript. AA and HS adapted the questionnaires and wrote the manuscript. SP analyzed and narrated the data. HAK and ARK edited and revised multiple drafts of the manuscript. ARK also assisted in adapting the questionnaires and posting it on social media channels of AKU. All authors reviewed and endorsed the final submission.

## Funding

The authors did not receive any direct funding for the purpose of this study.

## Acknowledgments

The authors would like to thank all the respondents of the survey from across Pakistan.

## Notes

### Competing Interest Statement

The authors have declared no competing interest.

### Clinical Trial

NA

